# Mapping plasmid–host networks and tracking persistent plasmids in wastewater microbiomes

**DOI:** 10.1101/2025.10.22.25338562

**Authors:** Siyi Zhou, Sarah E. Philo, Michael A. Saldana, Jeseth Delgado Vela, Adam L. Smith, Lauren B. Stadler

**Affiliations:** Civil and Environmental Engineering, Rice University, Houston, Texas, USA; Sonny Astani Department of Civil and Environmental Engineering, University of Southern California, Los Angeles, California, USA; Department of Civil and Environmental Engineering, Duke University, Durham, North Carolina, USA

**Keywords:** Plasmid-host associations, Water resource recovery facilities, Hi-C sequencing, Plasmid retention, Shotgun metagenomics

## Abstract

**Background:** Plasmids are major drivers of horizontal gene transfer (HGT), playing a central role in disseminating antimicrobial resistance in water resource recovery facilities (WRRFs). Most studies have focused on model or clinical plasmids in simplified settings, leaving their *in situ* dynamics in complex environmental communities poorly understood. To address this gap, we applied Hi-C metagenomic sequencing to systematically resolve plasmid–host associations and evaluate how environmental and operational factors influence plasmid persistence and host range across treatment stages and geographic regions in municipal WRRFs.

**Results:** We identified 944 plasmid clusters across influent, activated sludge, and effluent samples from three WRRFs located in three U.S. states, revealing distinct plasmid distribution patterns and plasmid-host associations across facilities and treatment stages. While overall bacterial community composition remained relatively stable across treatment stages, plasmid-host interactions varied, indicating that environmental conditions and treatment processes influenced plasmid retention and transfer. Plasmids in influent and activated sludge exhibited broader host ranges relative to effluent, where, plasmids were associated with a narrower set of bacterial hosts, likely reflecting the impacts of disinfection. Notably, certain plasmids persisted across treatment stages but exhibited substantial shifts in their associated bacterial hosts. Key taxa such as *Burkholderiaceae* and *Rhodocyclaceae* remained abundant throughout, indicating that shifts in plasmid-host associations were not solely driven by community turnover, but also suggesting HGT. Supporting this, functional profiling of effluent plasmids revealed enrichment in conjugation-related genes and virulence factors, including oxidative stress resistance, which may facilitate plasmid persistence and dissemination.

**Conclusions:** Our findings reveal that while wastewater microbial communities are diverse, plasmid-associated hosts are restricted to a few dominant bacterial families. Plasmid host range consistently narrows across treatment stages, reflecting the effectiveness of WRRFs in limiting plasmid persistence. However, shifts in host associations among highly similar plasmid clusters suggest potential HGT events. These results highlight the role of specific bacterial groups in plasmid dissemination and underscore the need for future studies to identify keystone taxa and mechanisms driving plasmid stability in complex microbial communities.

## Background

Plasmids are extrachromosomal genetic elements that facilitate bacterial adaptation and evolution by mediating horizontal gene transfer (HGT) [1–3]. These mobile genetic elements contribute to the dissemination of antimicrobial resistance (AMR), virulence factors, and metabolic capabilities, significantly shaping the function and dynamics of microbial communities [4–6]. While plasmid biology has been extensively studied under controlled laboratory conditions, the mechanisms governing plasmid persistence, such as conjugative transfer, fitness impacts, selective pressures, interspecies competition, remain to be assessed in complex environmental microbial communities [7–9]. Distinguishing between these mechanisms in representative environmental systems is critical for advancing our understanding of plasmid-host ecology.

Water resource recovery facilities (WRRFs) act as critical interfaces between the built and natural environment, facilitating the exchange and dissemination of genetic material, including AMR genes [9,10]. Biological treatment systems like activated sludge provide ideal conditions for HGT and the spread of adaptive traits due to high microbial densities, abundant biofilms, and active microbial metabolism [11–13]. Broad-host-range plasmids are key drivers of this process, transferring between diverse microbial hosts and mobilizing otherwise non-mobilizable plasmids, thereby accelerating the spread of AMR [6,14,15]. Although disinfection processes inactivate bacteria in wastewater effluents, they may not fully eliminate plasmid-carrying bacteria or extracellular DNA, allowing AMR genes to persist [16,17]. Residual plasmids in effluent have been reported as a concern, as they may contribute to the reintroduction of resistance genes into natural ecosystems, where they could be acquired by environmental or opportunistic pathogens [10]. Understanding plasmid persistence throughout treatment processes is crucial for assessing AMR risks and developing strategies to mitigate their dissemination from WRRFs to environmental microbial communities.

Despite advances in sequencing technologies, tracking plasmid dissemination and host range is methodologically challenging. Laboratory-based approaches, such as synthetic plasmid systems and culture-based conjugation assays, often rely on well-characterized, culturable hosts and controlled conditions, failing to capture the complexity of natural microbial ecosystems [18–20]. While emerging metagenomic approaches, including high-throughput single-cell sequencing, can be used to reveal host-MGE associations, they are limited by incomplete genome recovery, amplification biases, and insufficient sequencing depth, hindering the reconstruction of complete plasmid sequences and transmission networks [21,22].

Chromosome conformation capture sequencing, like Hi-C, preserves spatial interactions between plasmids and their hosts within intact microbial communities. This provides key ecological context for understanding plasmid dissemination in microbial communities [23–25]. Unlike single-cell sequencing, which isolates individual cells and may miss community interactions, Hi-C maintains *in situ* associations, enabling a more comprehensive reconstruction of plasmid-host networks. This capability is particularly valuable in complex environments such as WRRFs, where plasmids circulate among diverse taxa. Hi-C studies have inferred plasmid-host associations using either contig-level or bin-level data [24,25]. Contig-level analyses are highly sensitive, detecting links even for fragmented and low-abundance plasmids, but the loss of broader genomic context can inflate apparent host ranges. Bin-level analyses retain genomic context and improve host-range estimates, but they are more vulnerable to Hi-C noise. Artefactual links from contig ends or IS elements, as well as incomplete binning, can result in host misassignments [26]. In this study, to track plasmids from influent to effluent and examine their genetic context, we adapted a recent Hi-C-based plasmid clustering framework [23], and implemented a graph-based approach to group plasmid contigs into putative plasmid units with stringent linkage thresholds, aiming to provide a conservative, context-preserving basis for plasmid-host associations.

In this study, we applied Hi-C metagenomics and performed plasmid clustering to systematically track plasmid-host linkages across multiple unit processes within WRRFs in three U.S. states. Our objectives were to: (1) characterize the distribution of plasmids across treatment stages, (2) unravel plasmid-host associations with improved resolution of plasmid genetic context (e.g., encoded functions, plasmid mobility), and (3) identify key bacterial taxa that function as plasmid reservoirs. This study enhances our understanding of plasmid persistence and transmission in WRRFs by offering new insights into their microbial ecology.

## Methods

### Sample Collection and Concentration

Wastewater samples were collected weekly from three U.S. WRRFs in States A, B, and C, three times over a month. Primary influent was obtained as 24-hour composite samples (States A and B) or via grab sampling (State C). AS samples were collected via grab sampling from all sites, and effluent was grab-sampled after chlorination in States A and B, and combined UV/chlorination in State C. Effluent from State A underwent tertiary filtration to remove fine solids prior to chlorination. Samples from States A and C were shipped on ice to State B and stored at 4°C until processing. State A does not employ primary clarification; instead, influent undergoes preliminary screening and flows directly into the activated sludge process. State C operated a five-stage Bardenpho system with filtration and UV disinfection, while States A and B used high-purity oxygen activated sludge systems. All WRRFs discharged to local surface waters. Influent and AS samples were aliquoted into 50 mL and 2.0 mL tubes, respectively, centrifuged at 12,000 × g for 10 min at 4°C, and pellets stored at 4°C. Effluent (500 mL) was centrifuged at 3,500 × g for 30 min, pooled, recentrifuged, and pellets stored at 4°C. All samples were processed in duplicate: one replicate for Hi-C sequencing and the other for metagenomic DNA extraction.

### Hi-C sample preparation

One replicate per sample (by site, type, and date) was processed using the ProxiMeta™ Hi-C Kit (Phase Genomics, Seattle, WA, USA) following protocol v4.5.4, with modified crosslinking. Briefly, samples were incubated with 10 mL of chilled 1× PBS containing 1% formaldehyde for 10 min at room temperature, then quenched with 527 μL of 2.5 M glycine for 5 min. After a 10 min incubation on ice with agitation, samples were centrifuged at 3,500 × g for 10 min at 4°C, washed twice with cold 1× PBS, and resuspended in 1.0 mL PBS. A final spin at 800 × g for 5 min at 4°C was performed before storing the pellets at −80°C. Remaining steps followed the manufacturer’s instructions.

### DNA extraction and sequencing

Genomic DNA was extracted from pellet replicates using the Maxwell® RSC 48 system and Blood DNA Kit (Promega, Madison, WI, USA), with modified lysis to address solids in wastewater. Pellets were combined with 0.5 g zirconia beads (BioSpec) and 300 μL lysis buffer, bead-beaten at 2,400 RPM for 2 minutes, and centrifuged at 15,000 × g for 5 minutes. The supernatant (200 μL) was collected. This process was repeated twice with fresh lysis buffer, yielding 600 μL total lysate. Proteinase K (60 μL) was added, followed by incubation at 56°C for 30 minutes. DNA extracts were submitted to the Oklahoma Medical Research Foundation Clinical Genomics Core (Oklahoma City, OK) for library preparation using the Watchmaker DNA Library Prep Kit (Watchmaker Genomics, Boulder, CO) and sequenced on an Illumina NovaSeq X Plus (Illumina, San Diego, CA) to a depth of 100 million reads per sample. Indexed Hi-C libraries were pooled and sequenced separately at the Duke Sequencing and Genomic Technologies Core (Durham, NC) under the same platform and read depth.

### Shotgun metagenomic and Hi-C data analysis

Shotgun reads were quality-filtered and adapter-trimmed using BBDuk (BBTools, Joint Genome Institute) with the following parameters: k=23, ktrim=r, mink=12, hdist=1, minlength=50, –tpe, –tbo for adapter removal, and qtrim=rl, trimq=10, chastityfilter=True for low-quality base trimming. Trimmed reads were assembled using MEGAHIT with default parameters [27]. Shotgun and Hi-C reads were mapped to the assemblies using Bowtie2 and processed with Samtools, while Hi-C-specific alignments were performed using BWA-MEM [28–30]. Genome binning was conducted with MetaCC, which uses Hi-C contact frequencies to cluster contigs [31]. In total, binning across all samples recovered 988 metagenome-assembled genomes (MAGs) for State A, 1,475 MAGs for State B, and 882 MAGs for State C. Contact matrices were derived from Hi-C BAM files using the custom script *bam2links.py*, which quantifies linkage between contigs, as described by Press *et al* [32]. MAGs were annotated using Prokka, taxonomically classified with GTDB-Tk, and assessed for quality using CheckM [33–35]. While MAGs typically represent near-complete genomes of individual organisms, the quality of most MAGs in this study, as indicated by genome size and completeness estimates from CheckM, did not support this assumption. Accordingly, we interpret MAGs more broadly as genomes or genome fragments from closely related strains within a species. Plasmid contigs were identified and characterized using geNomad using the *end-to-end* command [36].

### Plasmid clusters identification and functional annotation

Putative plasmid contigs were identified using a Hi-C-based approach adapted from Risely *et al*.[23]. First, protein-coding genes were predicted from assembled contigs using Prodigal in metagenomic mode (-p meta) [37]. To minimize false-positive associations driven by mobile elements, contigs encoding transposases or insertion sequences (IS elements) were excluded. These were identified via BLASTp searches (e-value < 0.01) against curated transposase databases from ISfinder and the Tn3 Transposon Finder [38,39]. Contigs predicted to be plasmid-derived using geNomad were retained only if they exhibited strong linkage (≥15 Hi-C contacts) to at least one other plasmid contig, reflecting the assumption that genuine plasmids tend to co-occur stably in Hi-C networks. While this filtering may exclude small or non-transferable plasmids, it helps enrich for conjugative or mobilizable elements that are more ecologically relevant to HGT, as noted by Risely *et al*. [23]. Graph-based clustering of the retained contigs was then performed using the Walktrap algorithm (igraph::walktrap.community, step length = 10) [40], yielding a total of 944 plasmid clusters (Supplementary Fig. 1). Plasmid-cluster lengths ranged from 2,051 to 140,087 bp and were largely comparable across states (Supplementary Fig. 2). Lengths fall within reported ranges for natural plasmids [41,42]. Hi-C contact data were then integrated to identify host associations, revealing 289 plasmid clusters linked to at least one metagenome-assembled genome (MAG). These host-linked clusters were used in subsequent analyses. Hi-C contact counts were normalized for differences in host abundance and plasmid size, following the general approach of Risely *et al*. and Stalder *et al*. [23,25]. Specifically, for each MAG-plasmid pair, the raw Hi-C contact value was scaled by the average shotgun-derived MAG read count across all MAGs in the dataset relative to the read count for that MAG, and by the average plasmid cluster length across all plasmid clusters relative to the length of the individual plasmid cluster. This correction reduces detection biases introduced by variation in microbial abundance and plasmid size, enabling more comparable contact frequencies across samples (Supplementary Table 1).

Functional annotation was performed to target plasmid mobility, virulence factors, and AMR genes. MOB-suite (https://github.com/phac-nml/mob-suite) was used to classify plasmid clusters based on mobility-associated genes, including relaxases, origin-of-transfer (oriT) sites, and mating pair formation (MPF) systems [43]. Virulence factors were identified using ABRicate (https://github.com/tseemann/abricate) with the Virulence Factors Database (VFDB), retaining hits that met a minimum sequence identity of ≥75% and coverage of ≥70% [44]. AMR genes were detected using AMRFinderPlus, which identifies resistance determinants based on curated sequence models and HMM-based classification [45].

### Analysis of plasmid-host dynamics and plasmid persistence

To compare the total and plasmid-associated microbial communities, Hi-C and shotgun metagenomic data were aggregated across three weekly samples per stage. Relative abundances of bacterial families were used to assess community structure via non-metric multidimensional scaling (NMDS) based on Bray-Curtis dissimilarity (vegan::metaMDS) and evaluated by state and treatment stage using PERMANOVA (vegan::adonis2) [46]. Alpha diversity (Shannon index) was computed with vegan::diversity, and differences across groups were tested using Kruskal-Wallis and pairwise Wilcoxon tests (ggpubr::stat_compare_means) [47,48].

The number of plasmid clusters associated with each MAG and the host range of each plasmid cluster were quantified and compared across states. Plasmid-host bipartite networks were constructed from normalized Hi-C linkage data, retaining only associations with values ≥0.01 to exclude spurious contacts [25]. To evaluate plasmid persistence across wastewater treatment, we identified plasmid clusters that were highly similar (≥80% nucleotide identity) and detected across all three treatment stages. Sequence similarity between clusters was assessed using BLAST, and persistent plasmids were defined as those present in all three stages within a facility. Due to the fragmented nature of metagenomic assemblies and inherent variation in plasmid sizes, we did not apply a strict alignment length cutoff. While this approach may capture shared modular elements rather than entire plasmid sequences, it provides a conservative estimate of plasmid-associated sequence continuity across stages.

## Results

### Hi-C–derived plasmid-associated communities exhibit greater variability across states than total microbial communities

We began by comparing microbial and plasmid-associated communities across three states (A, B, and C) and three wastewater treatment stages: influent (Inf), activated sludge (AS), and effluent (Eff). Both were assessed at the MAG level (GTDB-Tk taxonomy), with plasmid-associated profiles including only MAGs linked to plasmids via Hi-C contacts. For each state and stage, the three weekly samples were combined and re-normalizing to produce a single representative community profile for that stage. Community-level variation was assessed using non-metric multidimensional scaling (NMDS) based on Bray–Curtis dissimilarities (Fig. 1a). Both shotgun and Hi-C–derived plasmid-host datasets revealed state-specific clustering of the microbial communities, with samples from the same state appearing more similar than those from different states. Furthermore, the Hi-C–based plasmid-associated communities showed more distant separation across states as compared to total communities. PERMANOVA analysis confirmed that geographical location (state) significantly influenced community composition (*p* = 0.015 for shotgun; *p* = 0.004 for Hi-C), whereas treatment stage had no significant effect (*p >* 0.9). Notably, the effect size was larger in the Hi-C dataset (R² = 0.508) than in the shotgun data (R² = 0.364), indicating that plasmid-associated communities were more variable across states than the total communities. This finding suggests that plasmid-host associations are more sensitive to differences in influent sources or treatment technologies across states than the overall microbial community structure.

**Fig. 1.**
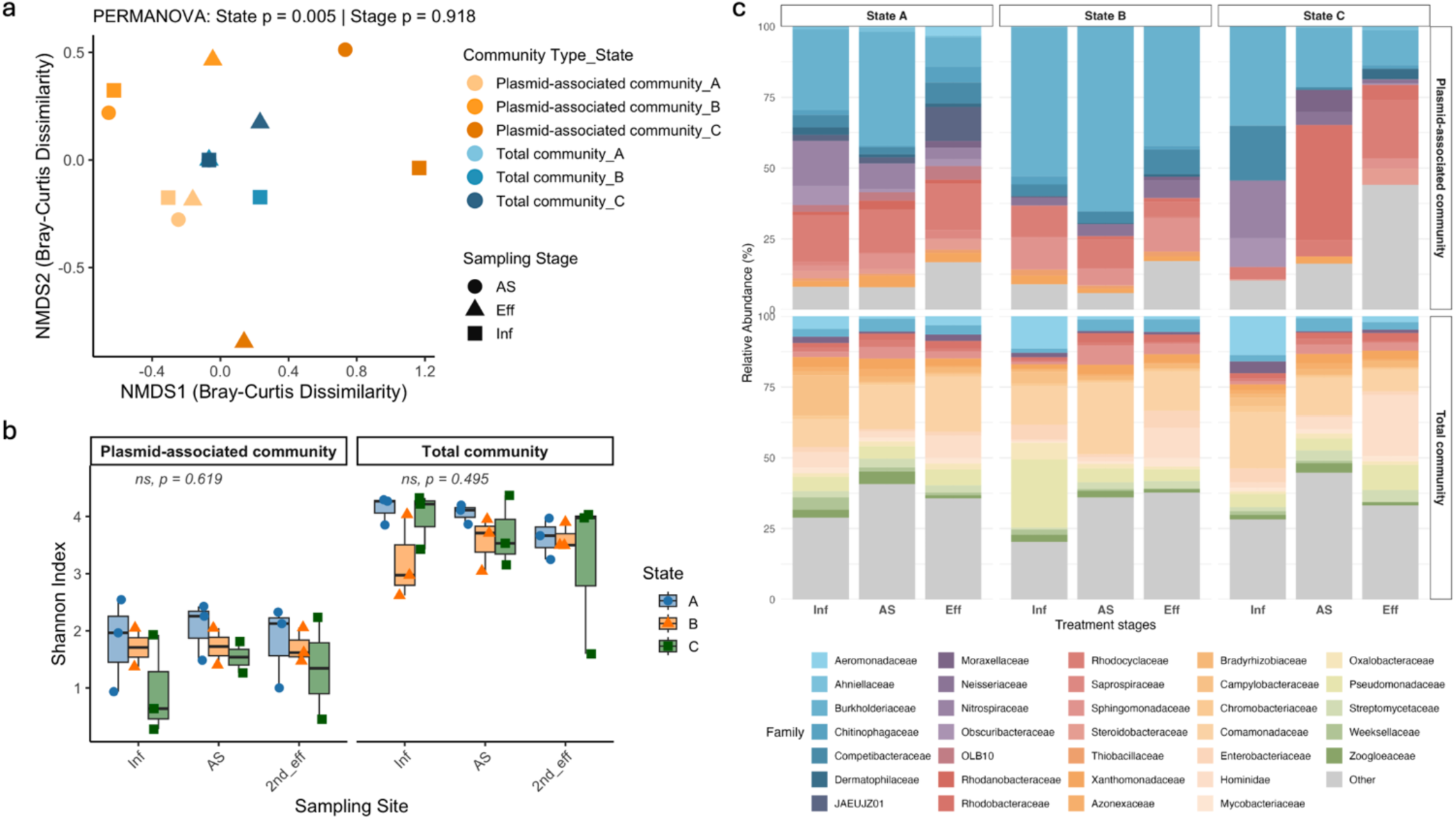
Microbial community composition, plasmid-associated taxa, and diversity patterns across states and wastewater treatment stages. (a) Non-metric multidimensional scaling (NMDS) plots based on Bray–Curtis dissimilarities show sample clustering by state for Hi-C-derived plasmid-associated taxa and shotgun-derived total bacterial communities. PERMANOVA revealed significant differences across states (Hi-C: *p* = 0.004, R² = 0.508; Shotgun: *p =* 0.015, R² = 0.364), while treatment stage had no significant effect (*p* > 0.9). (b) Shannon diversity (alpha diversity) of plasmid-associated (Hi-C) and total (shotgun) communities across samples. Shotgun data consistently showed higher diversity than Hi-C profiles, with no significant differences across states or stages (*p >* 0.05). (c) Relative abundance of the top 20 bacterial families in (top) plasmid-associated communities identified via Hi-C linkages to plasmid contigs and (bottom) total communities based on shotgun metagenomes. Taxonomic classifications are shown at the family level.

We next assessed alpha diversity using the Shannon index (Fig. 1b). Across all states and stages, total communities via shotgun metagenomes exhibited consistently higher diversity as compared to Hi-C-derived plasmid-associated communities. This reflects that plasmid-associated taxa represent only a subset of the overall microbial community. No significant differences in alpha diversity were observed across stages or states (*p* > 0.05), indicating that the diversity of total and plasmid-associated communities was relatively stable both across the treatment systems and across states. Additionally, Hi-C crosslinking may not capture all plasmid-bearing taxa, which could further contribute to lower observed diversity in the plasmid-associated fraction.

To better understand these differences in community composition between the total and plasmid-associated communities, we examined the taxonomic profiles derived from both shotgun and Hi-C data (Fig. 1c). We found that the dominant taxa were distinct between the total and plasmid-host communities. Specifically, shotgun metagenomes were dominated by *Comamonadaceae* (15.2% on average), followed by *Hominidae* (7.2%), *Pseudomonadaceae* (6.4%), *Aeromonadaceae* (4.6%), and *Burkholderiaceae* (4.4%). In contrast, the Hi-C-linked plasmid-associated taxa displayed a markedly narrower taxonomic spectrum. The dominant families in these profiles included *Burkholderiaceae* (34.3%), *UBA10799* (18.0%, within *Actinomycetes*), *Neisseriaceae* (15.3%), *Nitrospiraceae* (14.2%), and *Rhodocyclaceae* (12.6%). These findings highlight that the families that were most abundant in the overall community may not play major roles in plasmid retention. Instead, plasmid dissemination appears to be facilitated by a distinct set of bacterial families.

### Broader host range and higher plasmid prevalence in the State A WRRF may enhance HGT potential

To evaluate plasmid dissemination dynamics, we quantified: (1) plasmid host range, defined as either the number of unique metagenome-assembled genomes (MAGs; individual genome bins) or the number of distinct bacterial classes (taxonomic groups) associated with each plasmid cluster, and (2) plasmid burden, defined as the number of unique plasmid clusters associated with each MAG. Plasmids in the State A WRRF exhibited the broadest host ranges, averaging 8.8 ± 0.29 bacterial classes and 32.2 ± 1.48 MAGs per cluster, with some clusters linked to as many as 18 classes and 74 MAGs (Fig. 2a-b). In contrast, plasmids from the State B and State C WRRFs had lower host ranges, averaging 3.9 ± 0.25 and 3.4 ± 0.30 classes, respectively. These differences were statistically significant (p < 0.0001, Wilcoxon rank-sum test with Benjamini– Hochberg adjustment). Plasmid burden per MAG showed a similar trend (Fig. 2c; Supplementary Fig. 3): MAGs in the State A WRRF carried on average 18.6 ± 0.98 plasmid clusters (max = 57), compared to 10.5 ± 0.62 in State B (max = 46) and 5.1 ± 0.41 in State C (max = 29). These differences were also significant (p < 0.0001, Wilcoxon rank-sum test with Benjamini–Hochberg adjustment), indicating a higher plasmid load in the microbial community of the State A WRRF. The co-occurrence of broader plasmid dissemination and elevated plasmid burden may reflect enhanced HGT potential in the microbial communities of this facility.

**Fig. 2.**
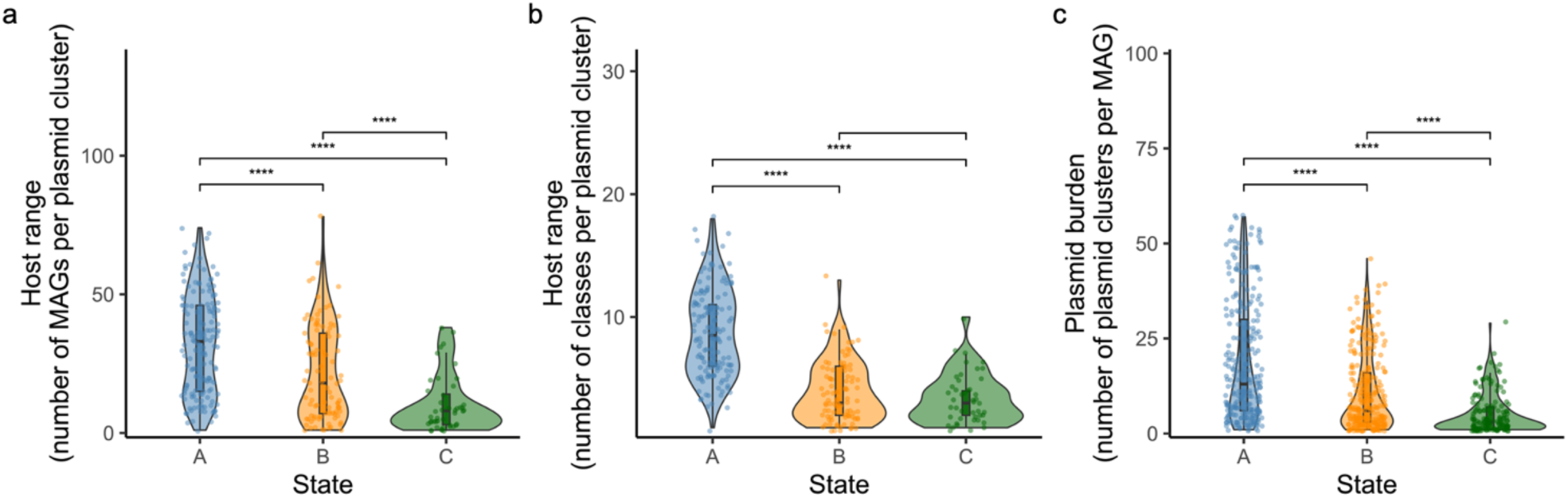
Plasmid-host associations and dissemination metrics across wastewater treatment samples. (a-b) Violin plots showing the distribution of plasmid host range, defined as the number of MAGs (a) and the number of bacterial classes (b) associated with each plasmid cluster in States A, B, and C. (c) Violin plot showing plasmid burden per MAG across states. For all panels, pairwise differences between states were evaluated using the unpaired Wilcoxon rank-sum test with Benjamini–Hochberg adjustment; significance levels are indicated by asterisks: *p* < 0.05 (*), *p* < 0.01 (**), *p* < 0.001 (***) and *p* < 0.0001 (****).

To further investigate WRRF-specific differences in plasmid-host interaction structure, we constructed bipartite networks linking plasmid clusters to bacterial host classes (Fig. 3a). The number of host-linked plasmid clusters varied among facilities (State A WRRF: 148; State B WRRF: 96; State C WRRF: 45), which may contribute to differences in apparent network complexity. Despite this, these networks revealed marked contrasts in connectivity across states. The State A WRRF showed a highly interconnected network, with many plasmid clusters linked to multiple host classes, indicating extensive cross-lineage dissemination. The State B WRRF displayed a more modular structure, with most plasmid clusters limited to *Gammaproteobacteria, Bacteroidia* and *Alphaproteobacteria*. The State C WRRF displayed sparse connectivity, suggesting a narrower host range and limited plasmid mobility within the microbial community.

**Fig. 3.**
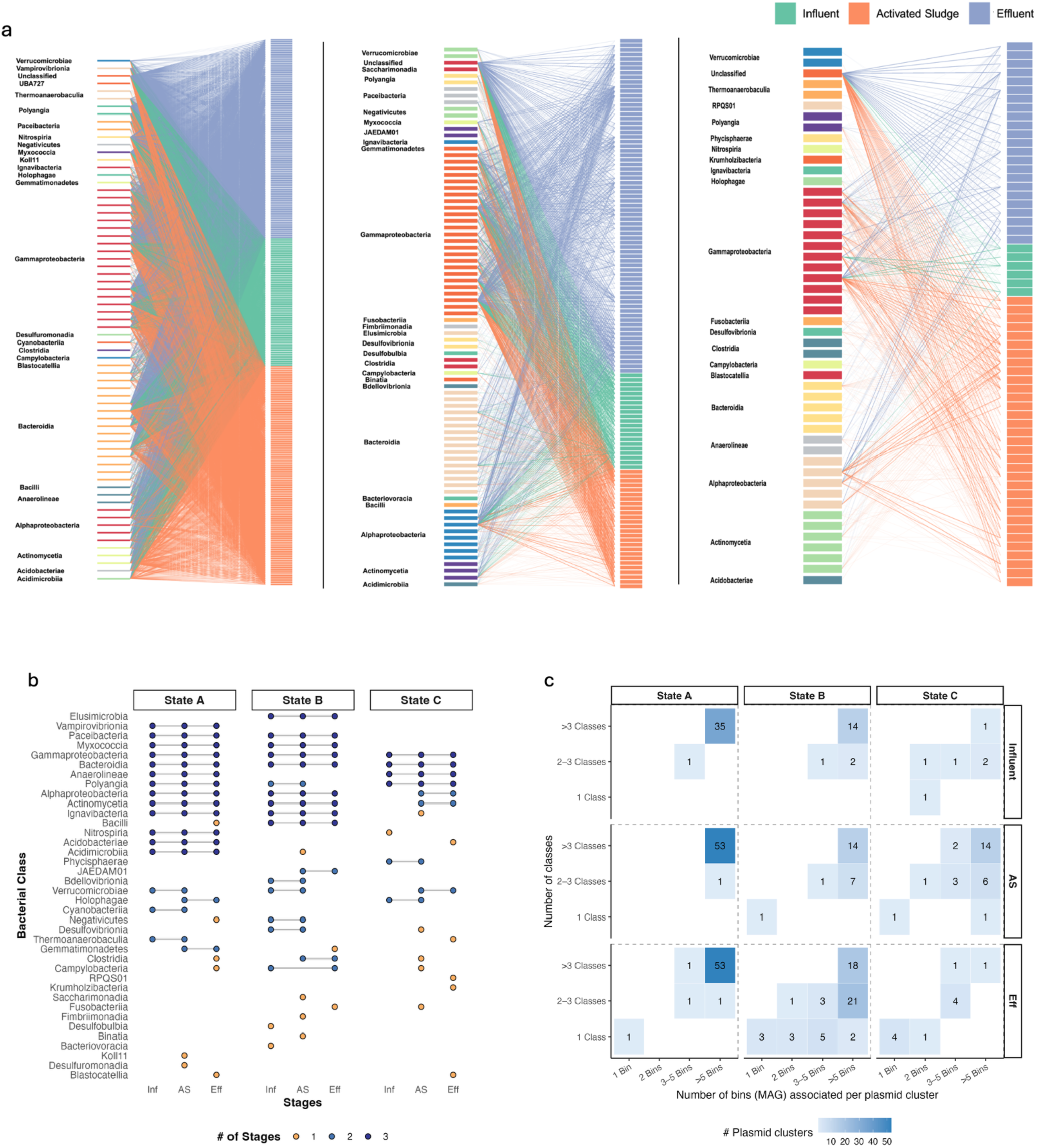
WRRF-specific patterns of plasmid-host association structure, host distribution across stages, and plasmid dissemination metrics. (a) Bipartite networks linking plasmid clusters (right columns, colored by treatment stages) to bacterial host families (left columns, colored by taxonomic class) in the State A, State B, and State C WRRFs. Edges represent plasmid–host associations inferred from Hi-C linkages. (b) Presence–absence matrix of plasmid-carrying bacterial classes across treatment stages. Each dot represents the detection of a given plasmid-carrying bacterial class. Dot color indicates persistence across stages: dark blue = detected in all three stages, light blue = detected in two stages, and yellow = detected in only one stage. (c) Heatmaps showing the number of plasmid clusters binned by host burden (i.e., the number of associated MAGs) and host range (i.e., the number of associated bacterial classes and bins) across treatment stages in each WRRF.

To explore the observed differences, we analyzed the persistence of plasmid-host associations across treatment stages within each state (Fig. 3b). The State A WRRF retained a diverse and stable set of host classes across all stages, with many associations maintained from influent to effluent. In contrast, the State B and State C WRRFs showed more stage-specific patterns, with fewer host classes shared between stages, indicating lower persistence of plasmid-host associations. We also quantified the occurrence of broadly distributed plasmids (e.g., ≥3 bacterial classes, ≥5 MAGs) across treatment stages (Fig. 3c). While such broad host ranges are relatively uncommon, cross-class transfer has been documented experimentally for promiscuous conjugative plasmids, including IncP-1, IncU, and IncW groups [49,50]. The State A WRRF consistently carried more broadly distributed plasmids, whereas most plasmids in the State C WRRF were confined to a single host class and a small number of MAGs. These findings suggest that the greater network connectivity observed in the State A WRRF is driven by broader plasmid host ranges and higher plasmid prevalence, reflecting greater HGT potential.

### Plasmid hosts shifted across treatment stages

We next investigated plasmid cluster distribution across treatment stages by quantifying their host range, defined as the number of associated MAGs and bacterial classes. Host range was calculated within each WRRF, and per-WRRF counts were pooled for stage-level comparisons. Plasmid host range differed significantly between stages: plasmids from influent and AS were associated with more MAGs and classes than those from effluent (*p* < 0.01, Wilcoxon test; Fig. 4a-b). WRRF-specific analyses (Supplementary Fig. 4a-b) showed a consistent overall pattern, though the degree of host range reduction from influent to effluent varied, likely reflecting differences in treatment configuration and/or influent composition. This suggests that plasmids with broader host ranges are more common in earlier stages of treatment, while those persisting in the effluent tend to be more host-restricted. Across all states, the number of MAGs per plasmid cluster was strongly correlated with the number of host classes per cluster (r > 0.81, *p* < 0.001; Fig. 4c), indicating that plasmids associating with more genomes also tend to span greater phylogenetic breadth.

**Fig. 4.**
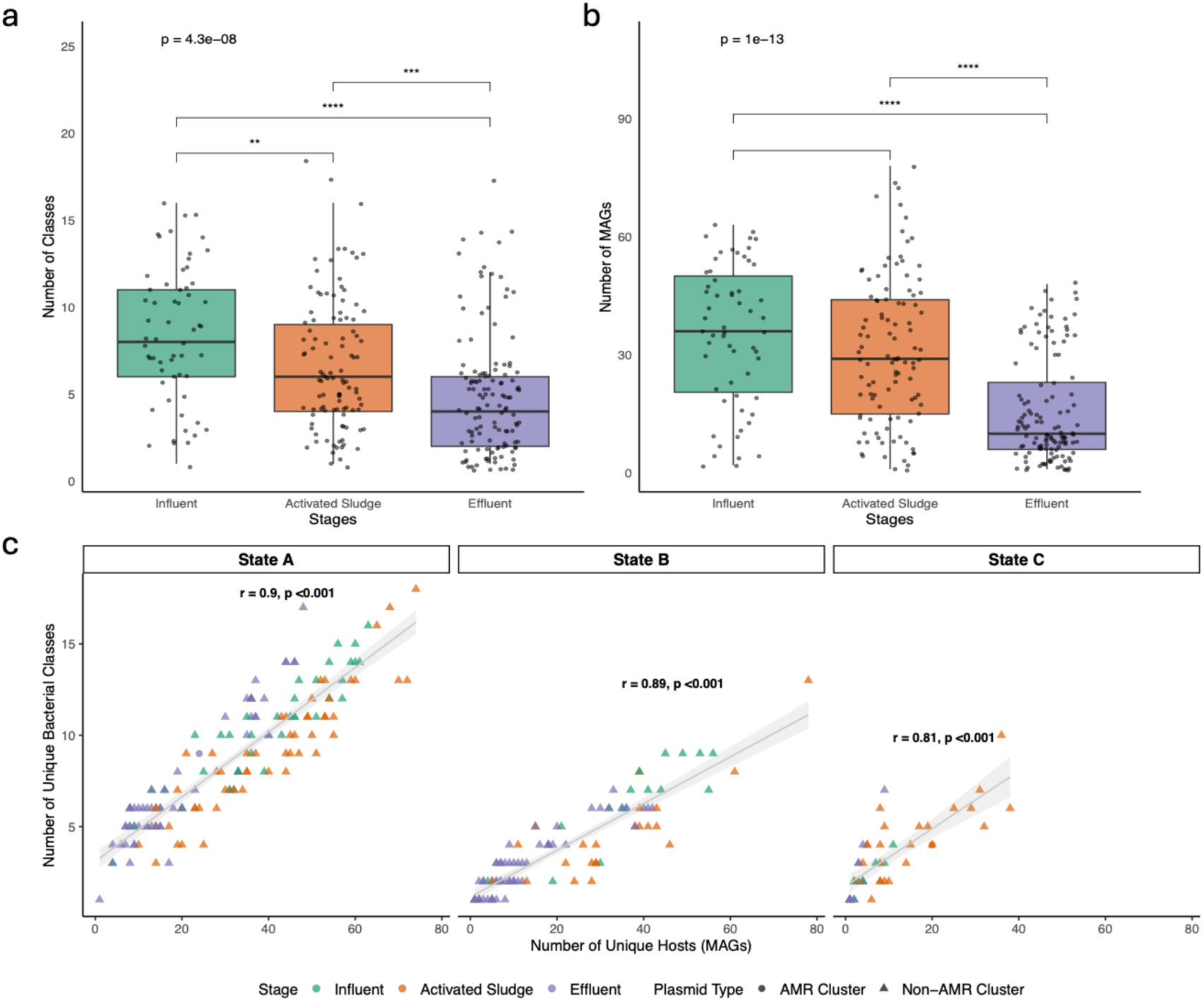
Plasmid host range across wastewater treatment stages. (a–b) Boxplots showing the distribution of host range metrics across stages: (a) number of unique bacterial classes and (b) number of unique MAGs per plasmid cluster. Asterisks indicate significance levels for pairwise comparisons: *p* < 0.05 (*), *p* < 0.01 (**), *p* < 0.001 (***) and *p* < 0.0001 (****). Dots represent individual clusters; box limits indicate interquartile range, with medians shown as bold lines. (c) Scatter plots showing the relationship between the number of unique MAGs (x-axis) and the number of unique bacterial classes (y-axis) associated with each plasmid cluster in the States A, B, and C WRRFs. Each point represents a plasmid cluster, colored by stage: influent (green), AS (orange), and effluent (purple). Spearman’s correlation coefficients (r) are shown for each state, with shaded areas indicating 95% confidence intervals.

To assess plasmid persistence across the wastewater treatment process, we then focused on structurally conserved plasmid clusters. Clusters were considered highly similar if they shared ≥80% nucleotide identity, representing conserved plasmid backbones. Plasmid clusters were deemed persistent if they were present in all three treatment stages from the same week. This analysis yielded 45 persistent plasmid clusters in the State A WRRF and 135 in the State B WRRF (Supplementary Table 2a-b). No persistent plasmid clusters were detected across multiple stages in the State C WRRF, likely due to limited detection of plasmid clusters in influent samples and their absence in several sampling timepoints, preventing cross-stage comparisons.

We next examined whether persistent plasmids maintained stable host associations or shifted hosts across the treatment process. We performed hierarchical clustering based on the host range of the persistent plasmid clusters and found that the plasmid clusters grouped mainly by treatment stage (Fig. 5 shows State A; Supplementary Fig. 5 shows State B). This pattern indicates that persistent plasmids frequently shift host associations between stages, suggesting dynamic host relationships and potential horizontal transfer during treatment. For instance, *Rhodocyclaceae* and *Burkholderiaceae* were frequent plasmid hosts in influent and activated sludge but not in effluent. Notably, their overall relative abundances remained stable across stages (Fig. 1c; Supplementary Fig. 6). This finding suggests that plasmid-host associations are not solely driven by host abundance but may instead reflect selective interactions or compatibility between specific plasmids and hosts in different environmental contexts.

**Fig. 5.**
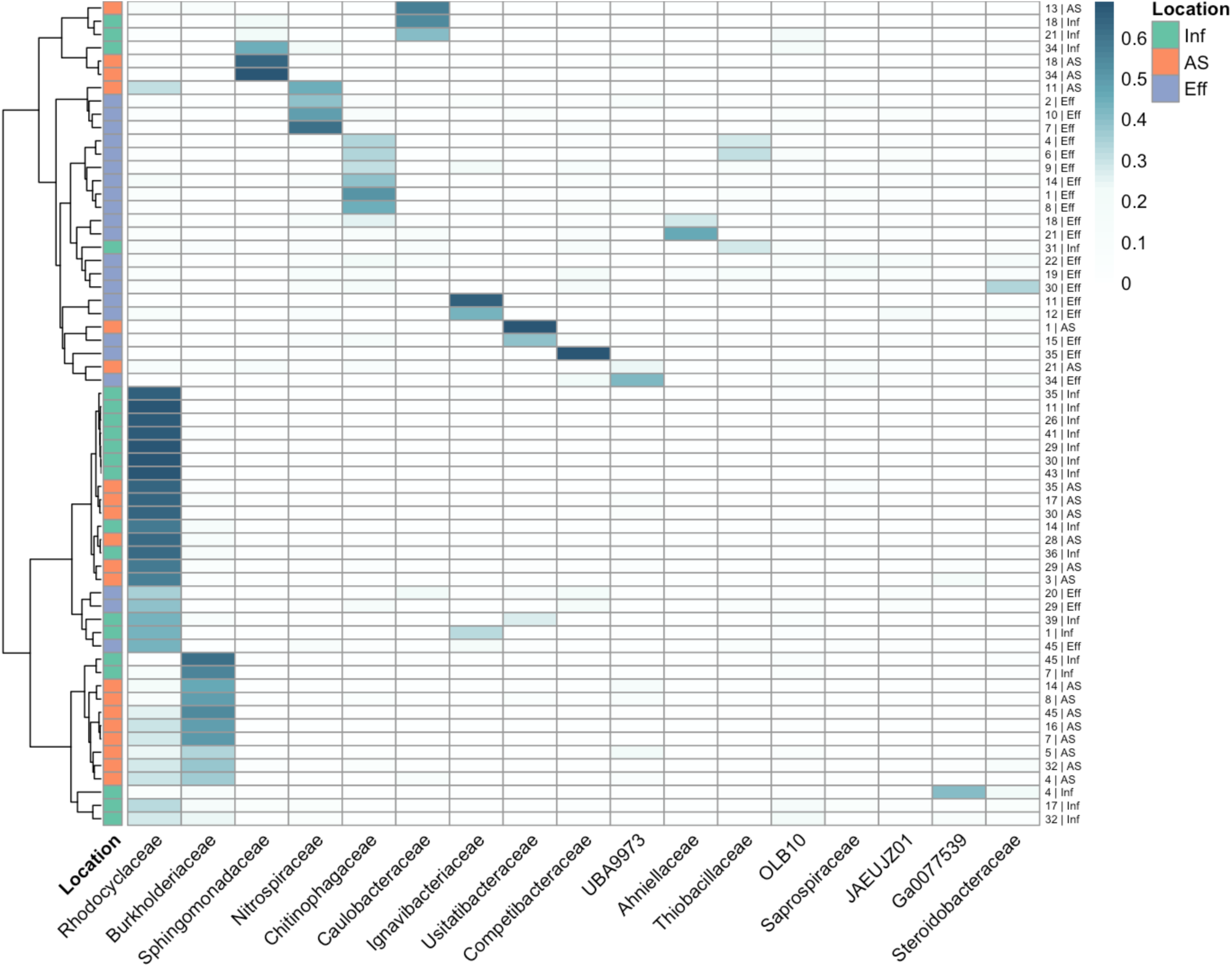
Shifts in host associations of persistent plasmid clusters across treatment stages. Heatmap for State A (State B in Supplementary Fig. 5). Columns are bacterial host families; rows are persistent plasmid clusters (≥80% sequence identity and detected in influent, activated sludge, and effluent within the same sampling week). Row labels are persistent ID | stage (Inf, AS, Eff). The right-side bar indicates treatment stage. Color intensity represents log-scaled normalized Hi-C contact frequency, reflecting the strength of plasmid-host associations. Rows are clustered by similarity in host-family profiles (y-axis dendrogram).

### Mobility potential and virulence content of plasmid clusters

We characterized the mobility potential of plasmid clusters by identifying genes involved in conjugation, including MOB genes and components of the Type IV secretion system (T4SS). A positive relationship was observed between the number of mobility-associated genes (MOB and T4SS) within plasmid clusters and the taxonomic breadth of their associated hosts (Supplementary Fig. 7a). Specifically, genes such as *MOBP*, *MOBQ*, *virB4*, and *trbC* were commonly detected in plasmid clusters linked to diverse host families, including members of *Gammaproteobacteria* (e.g., *Burkholderacace, Xanthomonacece, Competibactericaea*), *Actinobacteria* (e.g., *Mycobacteriaceae*), and *Bacteroidota* (e.g., *Chitinophageaceae*) (Supplementary Fig. 7b). In addition to conjugation genes, several plasmid clusters carried virulence factors (VFs) involved in oxidative stress defense (*HPI*, *KatG*), membrane remodeling (*PLD*), or resource acquisition (*HasD*). For example, one plasmid cluster from State B co-encoded the oxidative stress gene *HPI* along with multiple conjugation genes (*virB3*, *virB4*, *virB6*, *virB9*) and was linked to diverse host families (Supplementary Fig. 7c). These findings highlight a subset of potentially mobile virulence plasmids with broad host connectivity and provide additional support for the biological relevance and robustness of the identified plasmid clusters.

## Discussion

In this study, we evaluated shifts in plasmid-host associations across wastewater treatment stages in multiple states to understand plasmid persistence in complex microbial communities. We achieved this by adopting a recently developed methodology to group plasmid contigs into plasmid clusters. To improve upon contig-based limitations in host assignment and plasmid resolution, we applied a graph-based clustering approach that integrates Hi-C contact data with sequence similarity, grouping contigs into coherent plasmid clusters. This method reduces host association bias and allows for higher-resolution tracking of plasmid distribution and mobility potential across treatment stages using plasmid clusters. We also identified persistent plasmid clusters, defined as structurally conserved plasmid clusters present across all treatment stages, and used this approach to evaluate plasmid host shifts during wastewater treatment.

### Plasmid-host associations are selective and constrained to certain bacterial families

Across all WRRFs and treatment stages, plasmid-associated taxa constituted only a narrow subset of the total microbial community, indicating strong host–plasmid selectivity. Certain bacterial families, particularly *Burkholderiaceae*, *Nitrospiraceae*, and *Rhodocyclaceae*, were consistently overrepresented in plasmid-host networks, suggesting their enhanced permissiveness to plasmid acquisition and maintenance. These families harbor traits that likely support plasmid persistence. *Burkholderiaceae* are metabolically versatile and stress-tolerant, capable of producing antifungal compounds and withstanding oxidative stress, facilitating survival in dynamic environments [51,52]. *Nitrospiraceae*, including *Nitrospira marina*, exhibit redox adaptability and flexible energy metabolisms [53], while *Rhodocyclaceae*, such as *Azonexus*, are denitrifiers with broad substrate utilization, commonly found in wastewater systems [54]. These traits may facilitate their role as stable plasmid hosts in dynamic treatment environments. Plasmid persistence also depends on host fitness, conjugation rates, and interspecies competition [55,56]. Together, our findings suggest these families act as key plasmid reservoirs in wastewater microbiomes. Future work should identify and characterize plasmid-host pairs that consistently promote plasmid maintenance to elucidate the ecological and genetic factors stabilizing plasmids in complex communities.

### Operational and environmental context modulates plasmid host range and plasmid burden

Despite standardized sampling across treatment stages, plasmid prevalence and host range varied substantially across states. These differences likely reflect variation in operational design, influent characteristics, and environmental conditions. The WRRF in State A, which lacks primary clarification and uses a high-capacity activated sludge system with pure oxygen aeration, exhibited the broadest plasmid host range. This configuration supports high microbial biomass density and sustained aerobic activity, conditions favorable for conjugation and long-term plasmid maintenance [57–59]. Prior studies have shown that aeration and high dissolved oxygen concentrations can enhance plasmid host diversity by shaping microbial communities more permissive to HGT [60–62]. In contrast, the WRRF in State B also employs pure oxygen aeration but includes primary clarification. This step can selectively eliminate non-motile and floc-forming taxa via sedimentation, limiting the introduction of key bacterial groups into the aeration tanks and potentially reducing opportunities for plasmid transfer [63].

The WRRF in State C, which operates a five-stage Bardenpho process, exhibited the lowest overall plasmid host range and plasmid burden across stages. The alternating aerobic, anoxic, and anaerobic conditions inherent to this configuration likely impose energetic and regulatory constraints that limit plasmid stability. Previous work suggests that redox fluctuations can impair plasmid retention and horizontal transfer [64,65]. Additionally, frequent redox shifts, elevated biomass turnover, and rapid changes in microbial community composition may impose selective pressure against metabolically costly plasmids, narrowing host range and reducing plasmid permissiveness [66–68]. The relatively small catchment population served by the WRRF in State C may have also contributed to the lower observed diversity of incoming bacterial taxa and plasmids. These findings underscore the combined influence of redox regime and influent source diversity as key drivers of plasmid ecology in WRRFs, shaping both plasmid persistence and dissemination potential.

### Host switching and adaptive traits enable plasmid persistence through wastewater treatment

Although the overall bacterial community composition remained relatively stable across treatment stages, we observed a significant reduction in plasmid host range from influent to effluent, consistent with strong selective pressures imposed by wastewater treatment [69–71]. Persistent plasmids, identified across multiple treatment stages, were frequently associated with different host families at different points in the process. For example, the same plasmid backbones were linked to *Burkholderiaceae* in earlier stages but shifted to families such as *Thiobacillaceae* or *Nitrospiraceae* in the effluent. These shifts in host associations occurred even though the original host families remained abundant, suggesting that plasmid persistence is influenced more by plasmid-host compatibility and environmental selection than by host availability alone.

In fact, plasmid clusters spanning influent to effluent often encoded mobilization genes (e.g., *MOBQ, MOBP, MOBH*), type IV secretion system components, and adaptive traits such as oxidative stress resistance, membrane repair, and nutrient acquisition functions (Supplementary Fig. 5; Supplementary Table 2). These features likely enhance their ability to transfer and persist under treatment-imposed stressors like redox fluctuations and nutrient limitation. Previous studies similarly found that WRRFs act as ecological filters, favoring mobile and functionally adaptive plasmids while attenuating others [72,73]. For example, core class 1 integron gene cassettes with high resistance potential were found to persist through the treatment process, while others were attenuated, indicating selection against less stable genetic configurations [72].

Notably, we did not find any antibiotic resistance genes (ARGs) on persistent plasmids. This is likely because removing transposon- and IS-associated contigs excludes the highly dynamic regions where ARGs typically reside [6,23,26]. These findings underscore that both host dynamics and plasmid-intrinsic features contribute to the long-term stability and dissemination of plasmids in complex treatment environments.

### Methodological considerations and limitations

Our study has several limitations. First, Hi-C captures physical proximity, not definitive biological interaction, and may misassign hosts in dense microbial flocs despite conservative thresholds [23,25,26]. Second, plasmid recovery and clustering are limited by read depth and contig resolution, potentially underrepresenting low-abundance or cryptic plasmids. These factors likely differed across WRRFs and timepoints. As a result, the counts of host-linked plasmid clusters and the apparent network complexity may reflect sampling and recovery differences in addition to biological signal. We reduced these effects by normalizing Hi-C contacts to MAG-mapped reads and plasmid-cluster length, but detection and assembly biases remain. Excluding transposon- and IS-associated contigs further biases results toward more stable elements, possibly missing ARG-carrying plasmids [23]. Third, our study is based on DNA-level data. Without meta-transcriptomics or proteomics, we cannot determine the activity or expression of plasmid-borne genes, limiting functional inference [74]. Lastly, our sampling was limited to three states over three timepoints, which constrains statistical power and the generalizability of observed patterns. Broader spatiotemporal sampling would be needed to fully capture the variability in plasmid–host dynamics across WRRFs.

### Conclusions

This study provides new insights into plasmid–host associations in wastewater microbial communities and highlights that certain bacterial groups may play key roles in plasmid maintenance and dissemination. Although wastewater microbial communities are highly diverse, plasmid-associated hosts are constrained to a limited number of bacterial families. By sampling across three WRRFs in three different states, we showed that plasmid host range consistently narrowed from influent to activated sludge, and from activated sludge to effluent, reflecting the effectiveness of different WRRF designs in mitigating plasmid dissemination across treatment stages. These findings are consistent with previous studies suggesting that, although WRRFs have been considered potential “hot spots” for the spread of AMR, WRRFs are very effective at removing plasmids during treatment. Despite their ability to generally remove plasmids during treatment, certain plasmids are persistent across treatment. We found evidence that the hosts of persistent plasmids frequently shift during treatment. These findings suggest possible HGT events, although the underlying mechanisms require further study. Additional evidence for HGT includes the frequent detection of mobility genes and fitness-associated genes on persistent plasmids. Future studies should elucidate the mechanisms underlying plasmid stability in complex communities, which is critical to understanding and mitigating the environmental dissemination of AMR.

## Data Availability

All data produced in the present work are contained in the manuscript; All sequencing data generated in this study are deposited in the NCBI Sequence Read Archive (SRA) database under the BioProject ID: PRJNA1295499 (https://www.ncbi.nlm.nih.gov/bioproject/PRJNA1295499).

## List of abbreviations

AMR: Antimicrobial resistance
ARG: Antibiotic resistant genes
HGT: Horizontal gene transfer
ISs: Insertion sequences
WRRFs: Water resource recovery facilities
MGEs: Mobile genetic elements
MAGs: Metagenomic assembled genomes
VFs: Virulence factors

## Declarations

### Ethics approval and consent to participate

Not applicable

### Consent for publication

Not applicable

### Availability of data and materials

All sequencing data generated in this study are deposited in the NCBI Sequence Read Archive (SRA) database under the BioProject ID: PRJNA1295499 (https://www.ncbi.nlm.nih.gov/bioproject/PRJNA1295499). All code needed to reproduce the plasmid clustering and Hi-C plasmid–host linking analyses is available at https://doi.org/10.17605/OSF.IO/STMU9. Other relevant information is included in the manuscript and the supplementary files.

### Authors’ contributions

SZ designed the study, collected samples, conducted computational analyses, and drafted the manuscript. SEP and MAS contributed to sample collection, laboratory processing, and manuscript revision. JDV contributed to data interpretation and manuscript editing. ALS provided funding support and contributed to manuscript revision. LBS supervised the project, contributed to study design, and provided critical feedback. All authors reviewed and approved the final manuscript and agreed to be accountable for all aspects of the work.

## Acknowledgements

We thank the wastewater utilities for providing access to sampling sites. We also thank the participating wastewater treatment facilities for enabling sample collection.

