## Supplementary Figure 1 - 7 for "Mapping plasmid–host networks and tracking persistent plasmids in wastewater microbiomes"

### Supplementary Figures

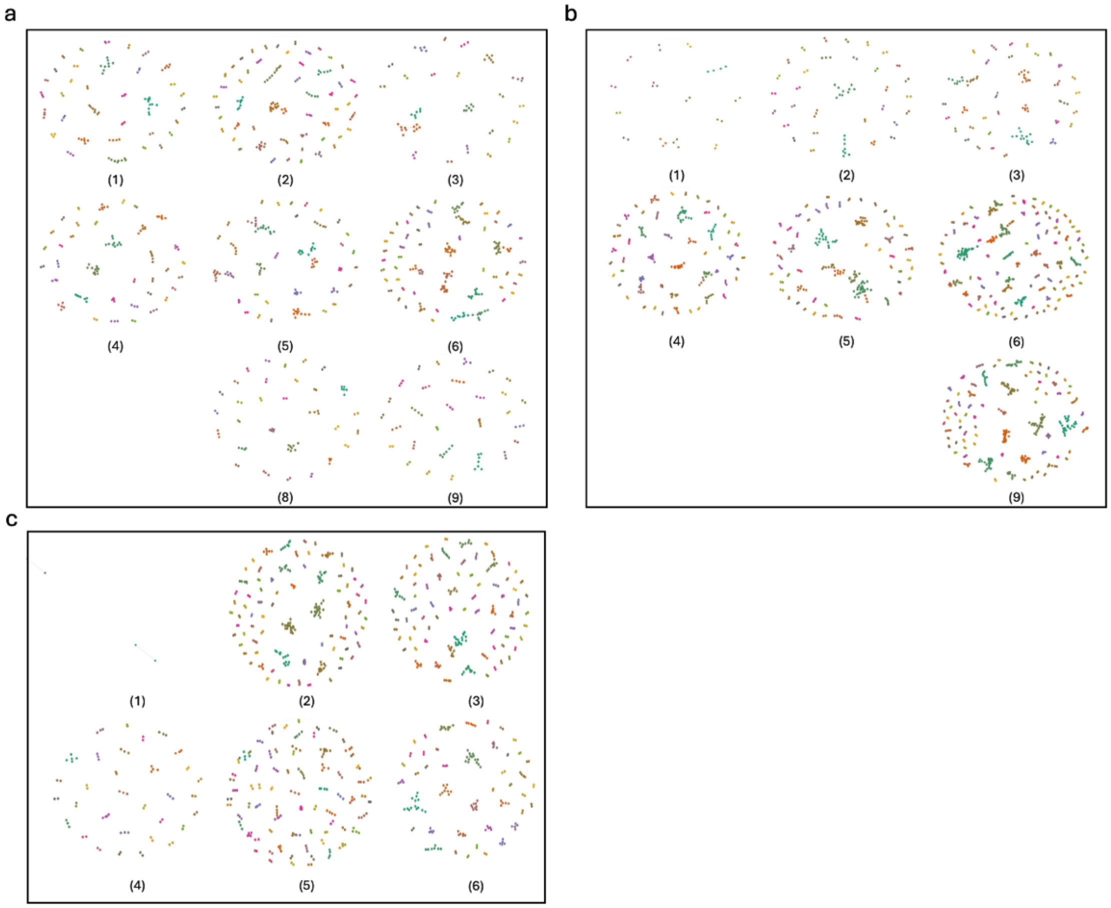


**Supplementary Figure 1.** Plasmid clusters detected in wastewater samples from three states, shown in panels (a-c) for states A, B, and C, respectively. Each dot represents a plasmid contig, and clusters were defined based on Hi-C linkages using the Walktrap algorithm. Each subplot is labeled (1-9), corresponding to samples collected across three weeks (top to bottom) and three treatment stages: Influent, Activated Sludge, and Effluent (left to right). Dotted outlines represent individual plasmid clusters. Samples (e.g., State A (7), B (7-8), C (7-9)) were omitted when their Hi-C libraries had insufficient usable read coverage and/or failed to meet the inter-plasmid linkage threshold (≥15 contacts to another plasmid contig), resulting in no recoverable clusters.

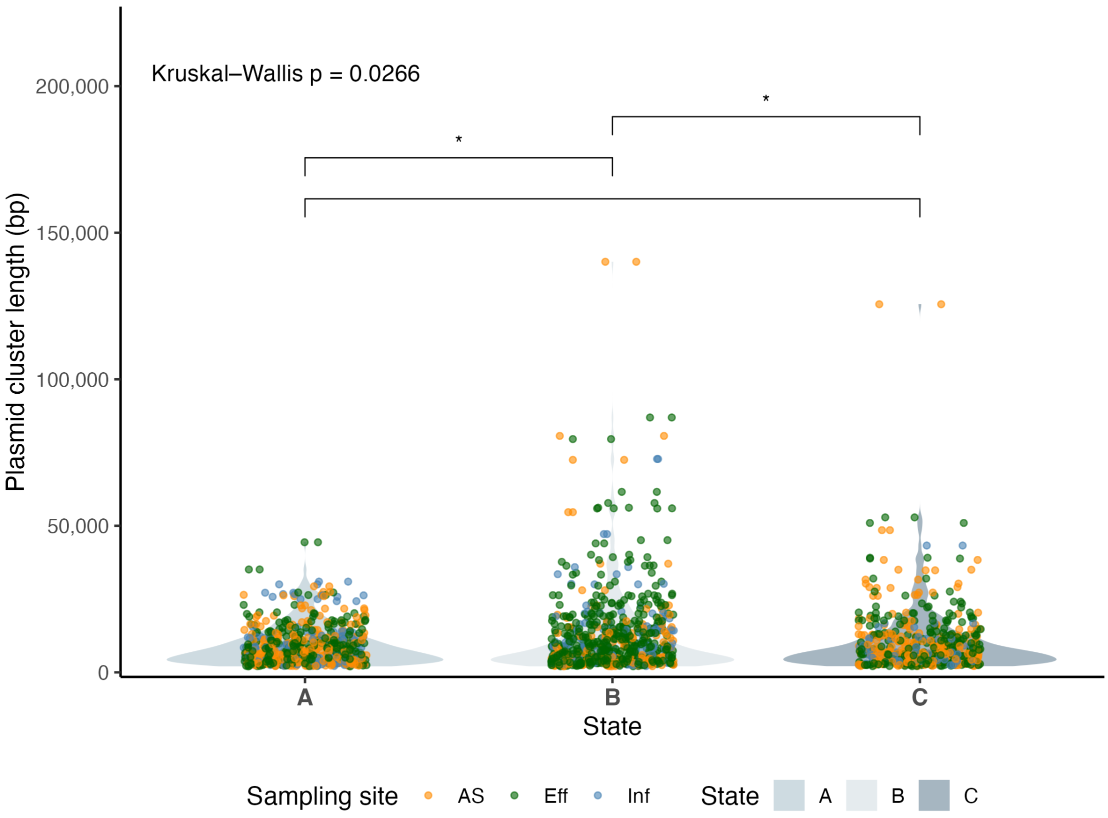


**Supplementary Figure 2.** Plasmid-cluster length distributions by state and treatment stage. Violin plots show cluster-length distributions for States A, B and C; points are individual clusters colored by stage (Influent, Activated Sludge, Effluent). Lengths were broadly comparable across states, with a modest overall difference (Kruskal–Wallis, p = 0.0266). Benjamini–Hochberg–adjusted pairwise Wilcoxon tests indicated B differed from A (p = 0.0356) and from C (p = 0.0356), whereas A vs C was not significant (p = 0.834).

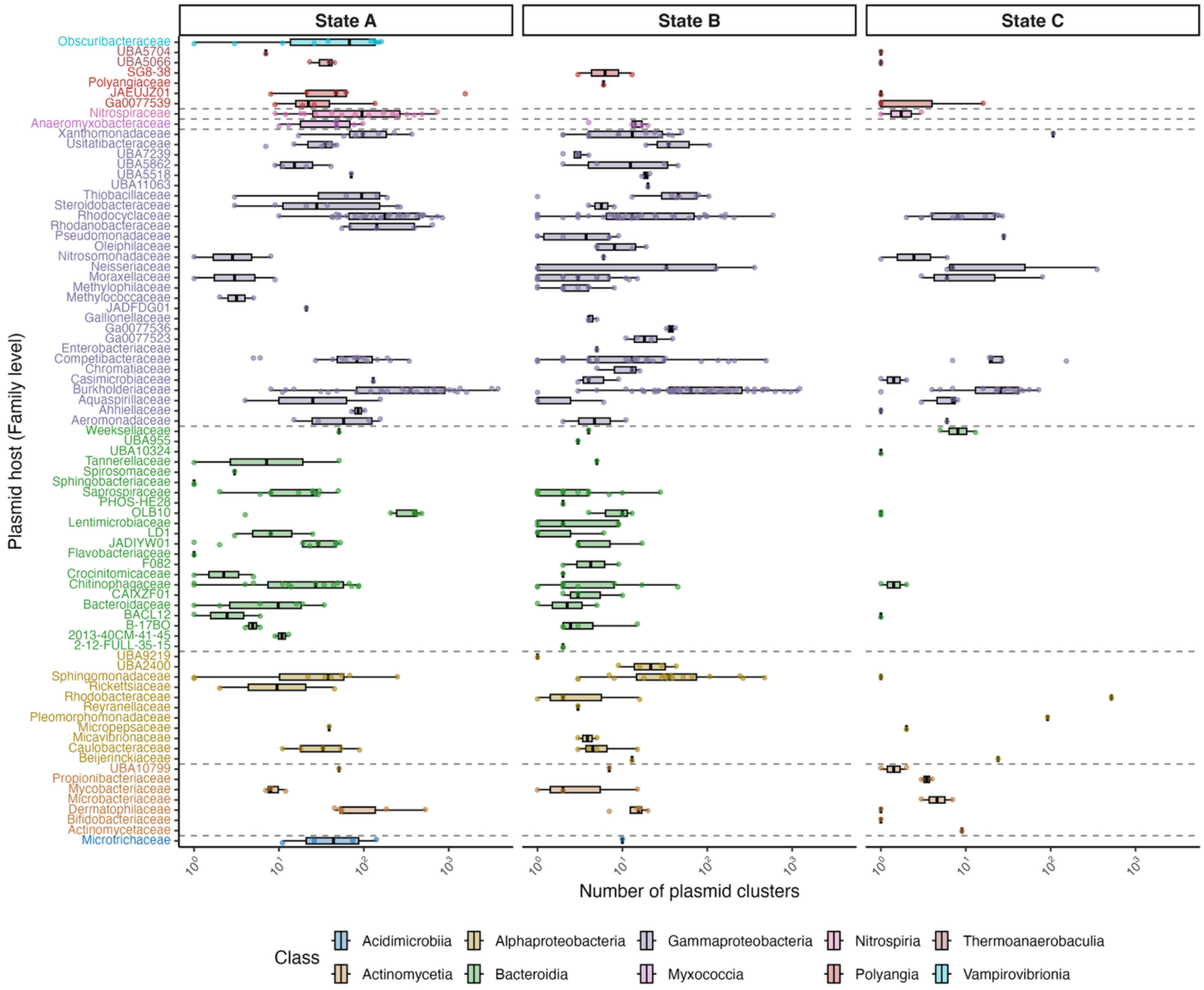


**Supplementary Figure 3.** Distribution of plasmid burden across bacterial families in three states. Plasmid burden is the number of plasmid clusters linked by Hi-C to each MAG. Each panel shows MAG-level plasmid burden for families detected in State A, State B, and State C wastewater samples. Points are individual MAGs; boxplots summarize per-family distributions. Families are grouped and color-coded by bacterial class. Horizontal dashed lines separate families belonging to different classes. The x-axis is log-scaled to reflect variation in plasmid burden. Only the top 10 most prevalent bacterial classes across all samples are highlighted.

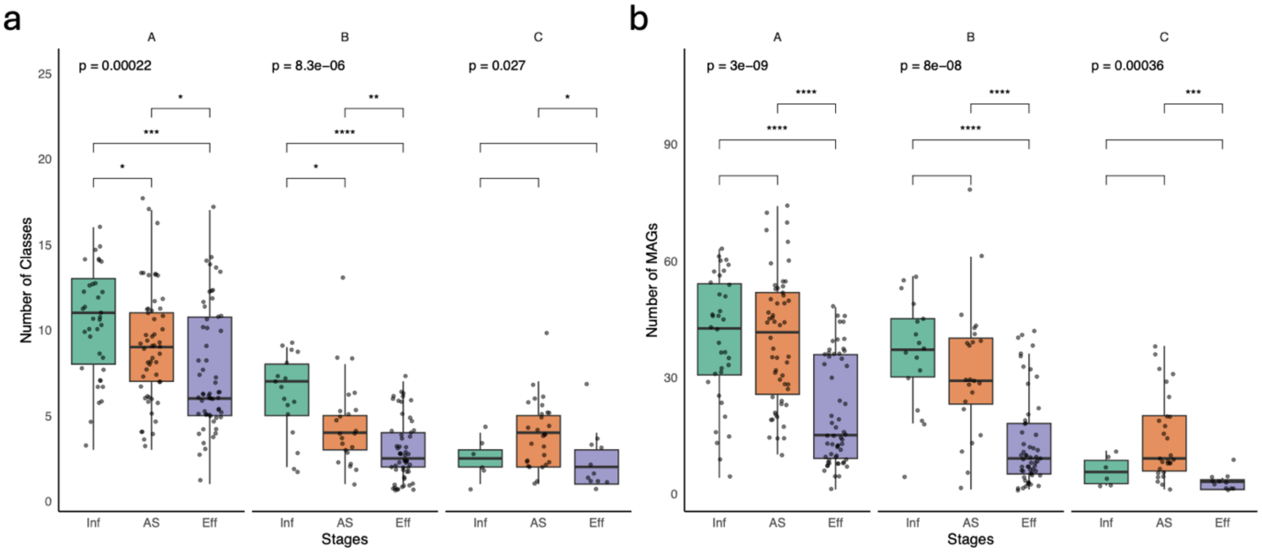


**Supplementary Figure 4.** Differences in plasmid host range across treatment stages in three WRRFs. (a) Number of bacterial classes linked to plasmid clusters per sample. (b) Number of MAGs linked to plasmid clusters per sample. Boxes show medians (lines), interquartile ranges (boxes), and 1.5× IQR whiskers; points represent individual samples. Colors indicate treatment stage: influent (green), activated sludge (orange), and effluent (purple). Statistical significance determined by pairwise Wilcoxon rank-sum tests; p values adjusted using the Benjamini–Hochberg method.

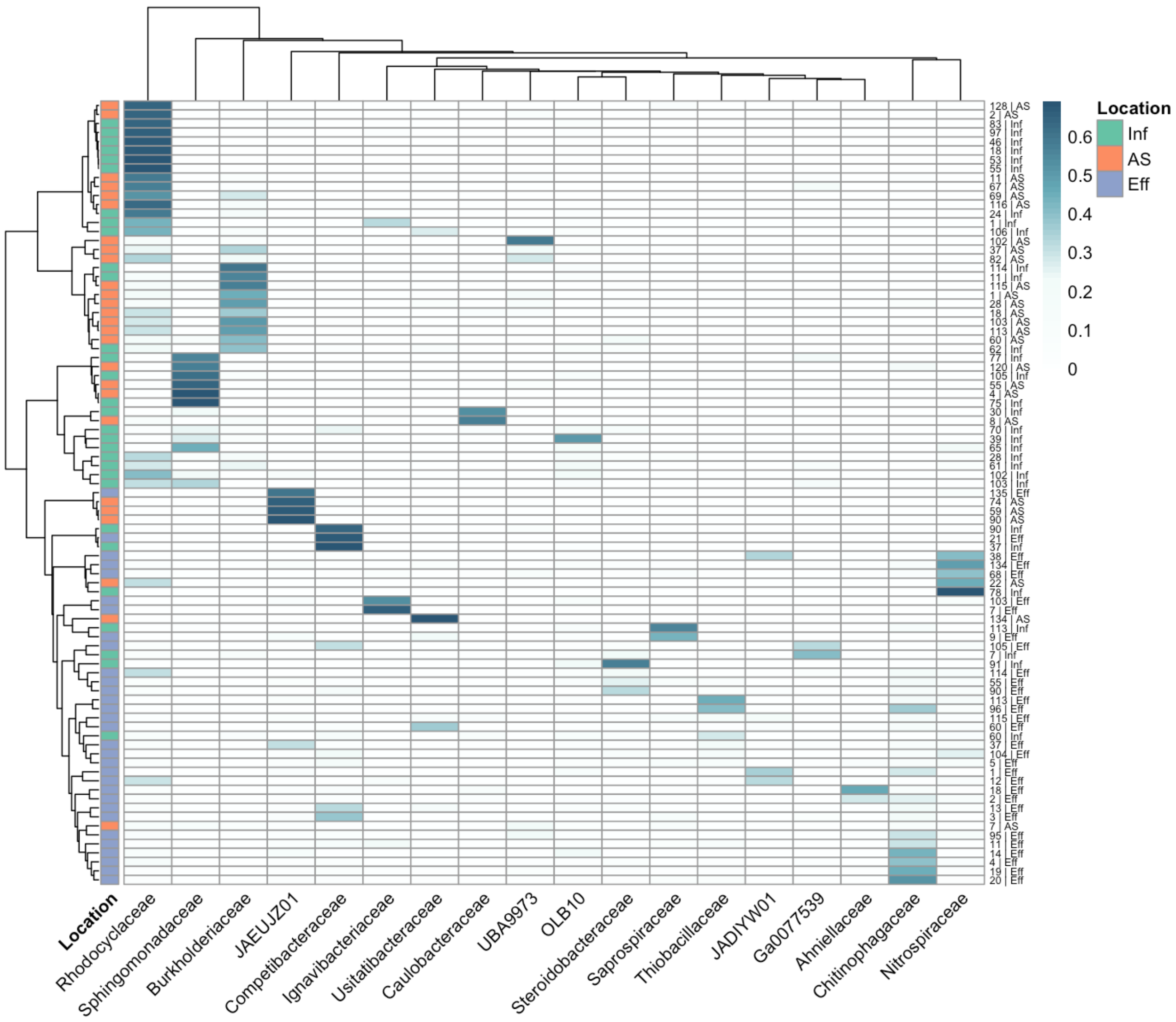


**Supplementary Figure 5.** Shifts in host associations of persistent plasmid clusters across treatment stages in State B. Columns are bacterial host families; rows are persistent plasmid clusters (≥80% sequence identity and detected in influent, activated sludge, and effluent within the same sampling week). Row labels are persistent ID | stage (Inf, AS, Eff). The right-side bar indicates treatment stage. Color intensity represents log-scaled normalized Hi-C contact frequency, reflecting the strength of plasmid-host associations. Rows are clustered by similarity in host-family profiles (y-axis dendrogram).

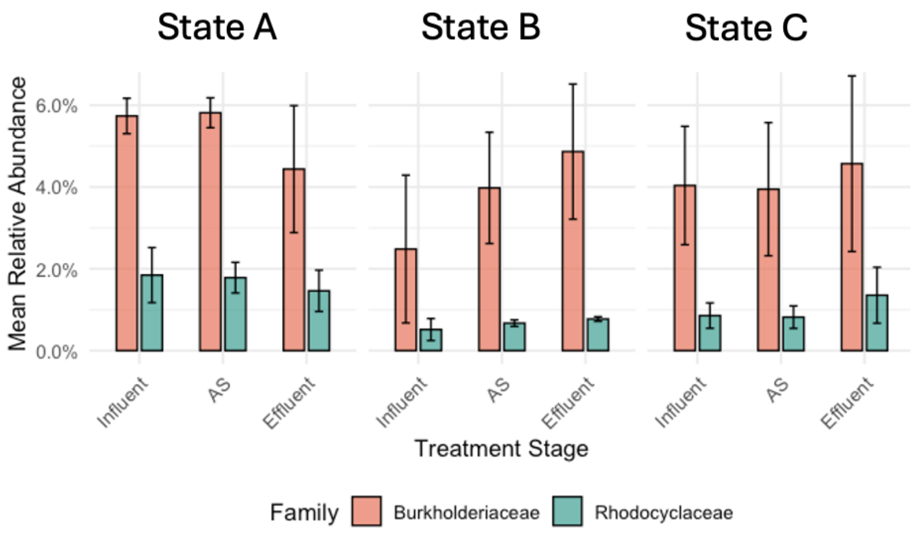


**Supplementary Figure 6.** Relative abundance of *Burkholderiaceae* and *Rhodocyclaceae* across wastewater treatment stages in three states. Bars represent the average relative abundance of each bacterial family in influent, activated sludge (AS), and effluent samples, grouped by state. Within each state, stage-wise differences were not significant for either family (Wilcoxon test with Benjamini–Hochberg correction; all pairwise $p > 0.05$).

a

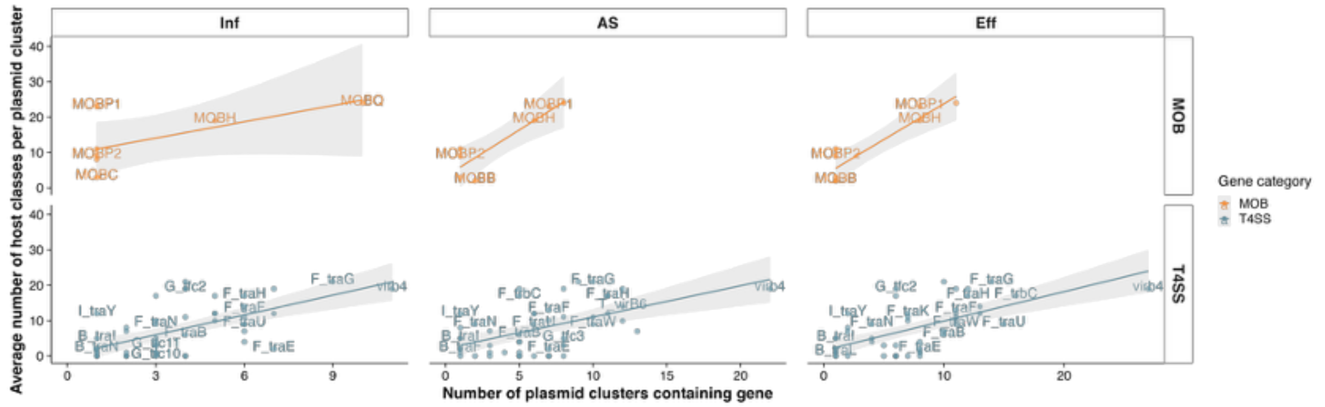

Inf
AS
Eff
MOB
T4SS
Average number of host classes per plasmid cluster
Number of plasmid clusters containing gene
Gene category
MOB
T4SS


b

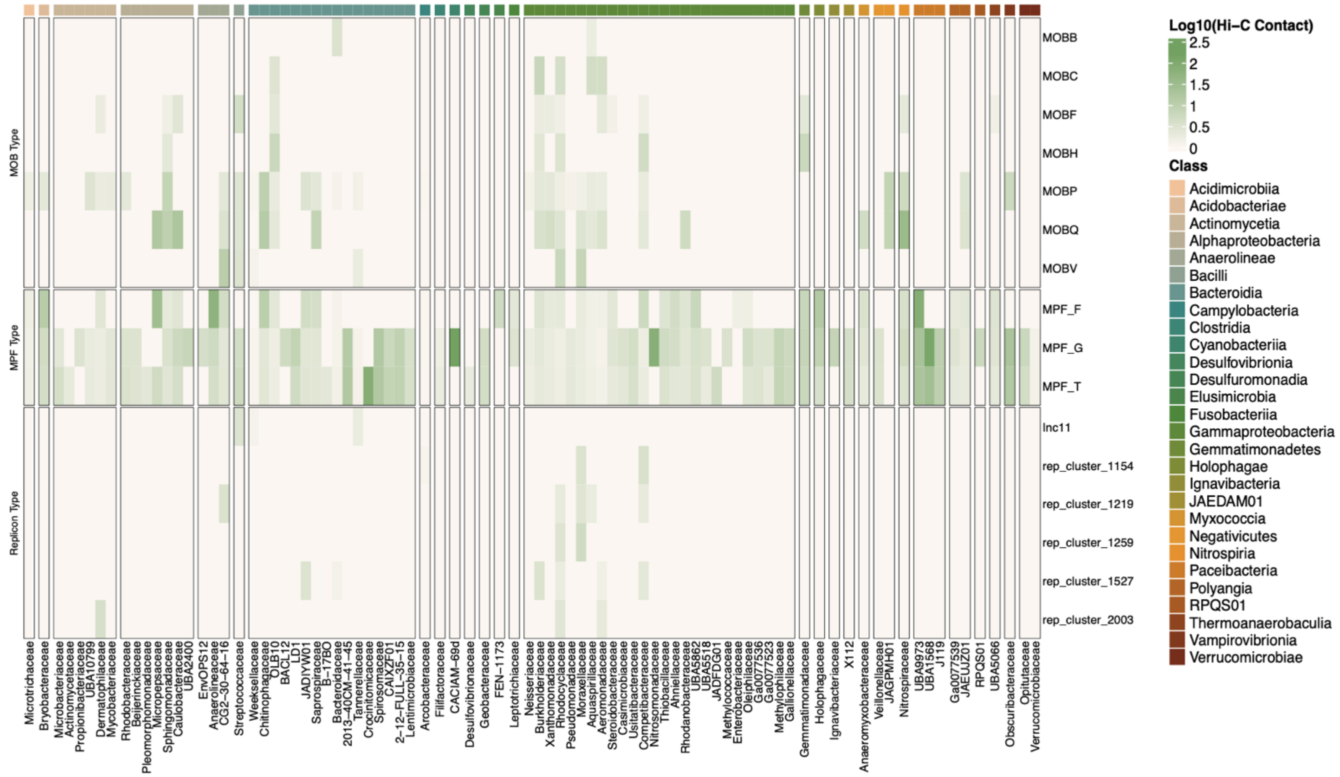

Log10(Hi-C Contact)
Class
Acidimicrobiia
Acidobacteriae
Actinomycetia
Alphaproteobacteria
Anaerolineae
Bacilli
Bacteroidia
Campylobacteria
Clostridia
Cyanobacteriia
Desulfovibrionia
Desulfuromonadia
Elusimicrobia
Fusobacteriia
Gammaproteobacteria
Gemmatimonadetes
Holophagae
Ignavibacteria
JAEDAM01
Myxococcia
Negativicutes
Nitrospiria
Paceibacteria
Polyangia
RPQS01
Thermoanaerobaculia
Vampirovibrionia
Verrucomicrobiae
MOB Type
MPF Type
Replicon Type
MOBB
MOBC
MOBF
MOBH
MOBP
MOBQ
MOBV
MPF_F
MPF_G
MPF_T
Inc11
rep_cluster_1154
rep_cluster_1219
rep_cluster_1259
rep_cluster_1527
rep_cluster_2003

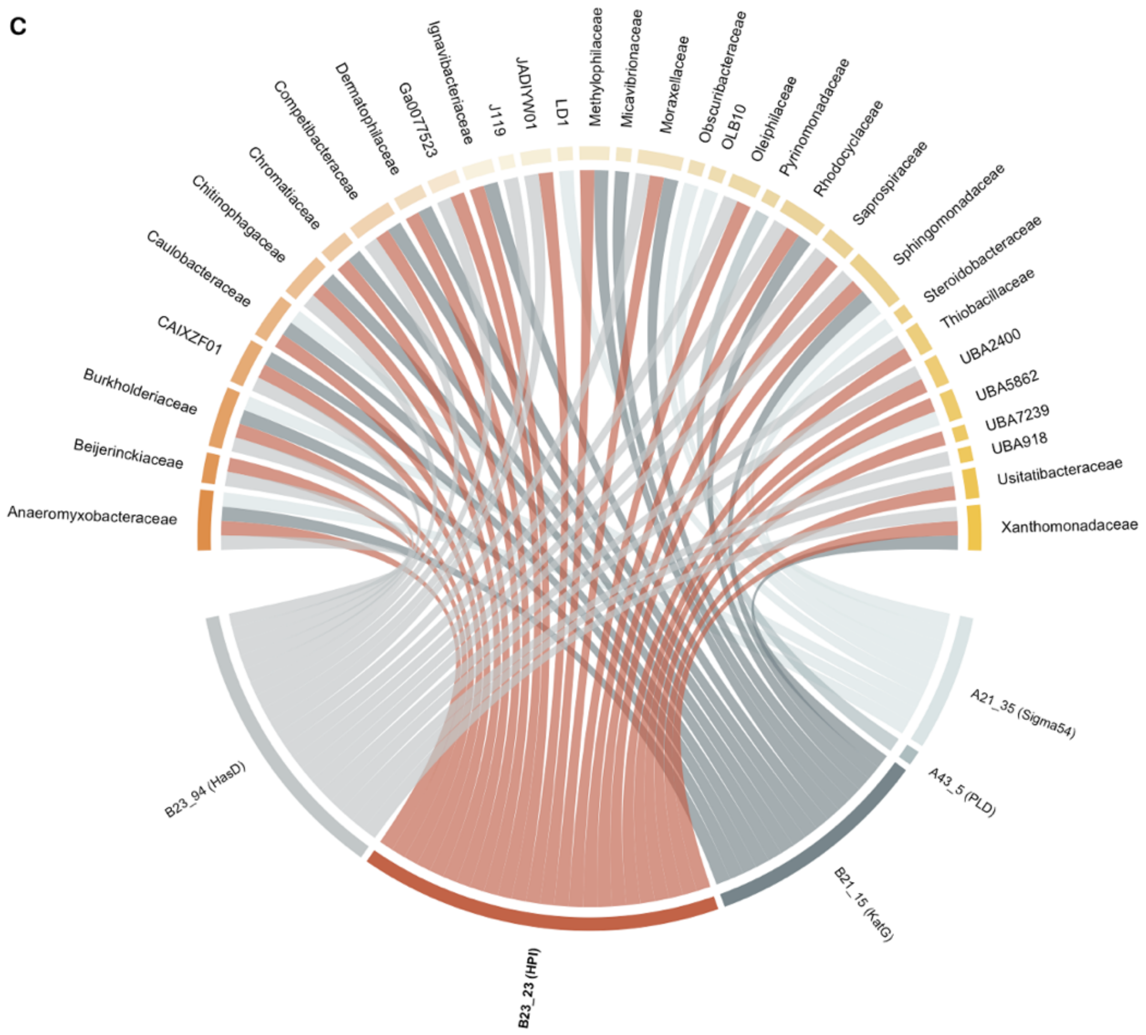


**Supplementary Figure 7.** Conjugation gene distribution, host range, and co-occurrence with virulence factors. (a) Scatterplots showing the relationship between the number of plasmid clusters carrying each conjugation gene and the average number of host classes associated with those clusters across compartments. Each point represents an individual gene; trend lines reflect linear regression with 95% confidence intervals. (b) Heatmap of normalized Hi-C contact frequencies between conjugation-associated genes (rows) and bacterial host families (columns), grouped by gene type and host class. (c) Chord diagram linking virulence factor (VF)–carrying plasmid clusters (bottom arc, labeled with cluster ID and VF) to their associated bacterial host families (upper arc). Plasmid clusters encoding conjugation genes are highlighted in red.

### Supplementary Tables

**Supplementary Table 1 - Summary of identified plasmid clusters and their associated functional annotations across states and treatment stages.**

Provided in a separate Excel spreadsheet.

**Supplementary Table 2a-b - Summary of structurally conserved persistent plasmid clusters across wastewater treatment stages (State A & B).**

Provided in a separate Excel spreadsheet.
